# Landing-Related Mechanisms Predominate in Anterior Cruciate Ligament Injuries Among Young Athletes Regardless of Contact

**DOI:** 10.64898/2026.02.04.26345606

**Authors:** Shinsuke Sakoda

## Abstract

**Context:** ACL injury prevention in young athletes has traditionally relied on a dichotomous classification of contact versus noncontact mechanisms; however, this framework may not adequately capture the movement processes underlying many injuries.

**Objective:** To classify ACL injury mechanisms in athletes aged ≤22 years with a specific focus on landing-related movements and to examine their associations with sport contact characteristics and age.

**Design:** Retrospective observational study.

**Setting:** Single sports injury clinic.

**Patients or Other Participants:** A total of 151 athletes aged ≤22 years (mean age, 17.7 ± 2.1 years) diagnosed with ACL injury between January 2017 and November 2025.

**Main Outcome Measure(s):** ACL injury mechanisms were classified as landing-related without contact (L), landing-related with contact (Lc), or direct contact injury (C). Landing-related injuries (L + Lc) were compared with direct contact injuries. Multivariable logistic regression was used to identify factors associated with landing-related injury.

**Results:** Landing-related injuries accounted for 123 cases (81.5%), including 88 noncontact and 35 contact-related landing injuries, whereas direct contact injuries occurred in 24 cases (15.9%). Athletes with direct contact injuries were significantly older than those with landing-related injuries (19.2 ± 1.7 vs 17.5 ± 2.5 years, p = 0.03). In multivariable analysis, participation in noncollision sports was strongly associated with landing-related injury (odds ratio [OR] = 9.80; 95% confidence interval [CI] = 3.03–31.5; p < 0.001), whereas increasing age was inversely associated with landing-related injury (OR per year = 0.71; 95% CI = 0.56–0.90). Sex was not independently associated with injury mechanism.

**Conclusions:** Most ACL injuries in athletes aged ≤22 years occurred through landing-related mechanisms, regardless of contact. These findings highlight insufficient control of landing and foot contact as a fundamental mechanism of ACL injury in young athletes and support prevention strategies focused on movement quality during sport-specific tasks.

**Key Points:**

▪ Most ACL injuries in athletes aged ≤22 years occurred during landing or foot contact, regardless of whether external contact was present.
▪ Noncollision sports and younger age were strongly associated with landing-related ACL injury mechanisms.
▪ ACL injury prevention in young athletes should prioritize improving landing and foot contact control rather than focusing solely on contact characteristics.

## Introduction

Anterior cruciate ligament (ACL) injury is one of the most common and severe sports injuries among young athletes and is known to be associated with delayed return to sport, decreased athletic performance, and an increased risk of developing knee osteoarthritis later in life. ^1–3^ Accordingly, the prevention of ACL injury has been recognized as a major issue in the fields of sports medicine and athletic training. ^1^

Traditionally, ACL injury mechanisms have been broadly classified into contact and non-contact injuries. ^4^ Non-contact injuries are defined as those occurring without direct external force applied to the knee, and intrinsic factors such as impaired neuromuscular control, lower limb alignment, and movement patterns have been discussed as major contributors. ^1, 8, 9^ In contrast, contact injuries are attributed to direct external forces applied to the knee during player-to-player contact, and numerous epidemiological studies have focused on sport characteristics and contact frequency. ^4, 6, 7^

However, this dichotomous classification may not adequately reflect the actual injury situations that occur during sports activities in young athletes. ^5, 6^ In clinical practice, many ACL injuries occur during jump landing or cutting maneuvers without obvious direct external force applied to the knee, and injuries also frequently occur during landing movements immediately after player contact. ^4–7^ These observations indicate that organizing ACL injury mechanisms solely based on the presence or absence of contact has inherent limitations. Furthermore, although recent ACL injury prevention strategies have increasingly focused on neuromuscular training and improvements in movement patterns, many prevention programs continue to treat noncontact injuries as a single category, and classification based on specific movement processes such as landing and foot contact remains insufficient. ^1, 5^ This limitation may be particularly relevant in young athletes, in whom differences in movement control related to growth and developmental stage can influence injury mechanisms. ^10^

Therefore, in this study, ACL injuries occurring during jump landing or foot contact, with or without preceding player contact, were collectively defined as landing-related injury mechanisms, and their characteristics were explored. The purpose of this study was to classify ACL injury mechanisms in athletes aged 22 years or younger, with a specific focus on landing-related movements, and to examine their associations with sport contact characteristics and age.

## Methods

### Study Design and Participants

This retrospective observational study included patients aged 22 years or younger who were diagnosed with anterior cruciate ligament (ACL) injury and visited the sports injury clinic at our institution between January 2017 and November 2025. Eligible cases were extracted from an institutional sports injury database.

The inclusion criteria were a diagnosis of ACL injury based on clinical findings and/or imaging studies and the availability of detailed medical records describing the injury situation. Patients with insufficient information regarding the injury mechanism were excluded.

### Definition of Injury Mechanism

In this study, the term landing was used broadly to include not only landing from an airborne phase but also the moment of foot contact during cutting, planting, or deceleration, in which the movement vector changes abruptly.

ACL injury mechanisms were classified into three groups: landing-related injury without contact (L), landing-related injury with contact (Lc), and direct contact injury (C). The L group was defined as ACL injuries occurring without external contact during landing or foot contact, including cutting, deceleration, and planting movements.

The Lc group was defined as ACL injuries occurring during landing or foot contact following external contact that disrupted balance or posture.

The C group was defined as ACL injuries caused by direct external forces applied to the knee, such as valgus, rotational, or anterior–posterior forces during player contact.

Because both the L and Lc groups shared a common injury mechanism characterized by ACL injury occurring during landing or foot contact rather than from direct external forces applied to the knee, these groups were combined for analysis as the landing-related injury group and compared with the C group. Cases classified as other mechanisms were excluded from comparative analyses.

### Data Collection

From medical records, data on age at injury, sex, sport at the time of injury, and injury mechanism were collected.

Sports were categorized according to contact characteristics. Sports in which player-to-player collision is an inherent characteristic, such as rugby and combat sports, were classified as collision sports, whereas all other sports were classified as non-collision sports.

The injury mechanism was determined based on a detailed patient interview conducted at the initial visit. When available, video recordings or information from third parties were also reviewed. Classification into the L, Lc, or C groups was performed by the attending physician at the time of database registration according to predefined criteria.

### Outcome Measures

The primary outcome was the classification of injury mechanism, dichotomized as landing-related injury (L + Lc) or direct contact injury (C).

Secondary variables included age, sex, and sport contact characteristics, which were compared between injury mechanism groups.

### Statistical Analysis

Continuous variables were presented as mean ± standard deviation and were compared using Student’s t test or the Mann–Whitney U test, as appropriate. Categorical variables were presented as frequencies and percentages and were compared using the chi-square test or Fisher’s exact test.

To identify factors associated with landing-related injury, multivariable logistic regression analysis was performed. Age, sex, and sport contact characteristics were included as explanatory variables based on clinical relevance and previous literature. Results were presented as odds ratios (ORs) with 95% confidence intervals (CIs). Statistical significance was set at p < 0.05.

All statistical analyses were performed using R software (version 4.3.2; The R Foundation for Statistical Computing, Vienna, Austria).

### Ethics Approval

This study was approved by the institutional ethics committee of our institution. The requirement for informed consent was waived because of the retrospective study design.

## Results

### Participant Characteristics

A total of 151 patients aged 22 years or younger with anterior cruciate ligament (ACL) injuries were included in this study. The mean age was 17.7 ± 2.1 years. Of these patients, 84 (55.6%) were male and 67 (44.4%) were female.

### Distribution of Injury Mechanisms

Injury mechanisms were classified as noncontact landing-related injuries (L group) in 88 patients (58.3%), landing-related injuries following contact (Lc group) in 35 patients (23.2%), and direct contact injuries (C group) in 24 patients (15.9%). Other injury mechanisms were identified in 4 patients (2.6%).

When the L and Lc groups were combined, landing-related injuries (L + Lc) accounted for 123 cases (81.5%). Cases classified as other mechanisms were excluded from comparative analyses.

### Comparison Between Landing-Related and Direct Contact Injuries

Patients in the direct contact injury group were significantly older than those in the landing-related injury group (19.2 ± 1.7 years vs 17.5 ± 2.5 years, p = 0.03).

Sex distribution differed significantly between the groups, with a higher proportion of male patients in the direct contact injury group (p = 0.003, Table 1).

**Table 1.**
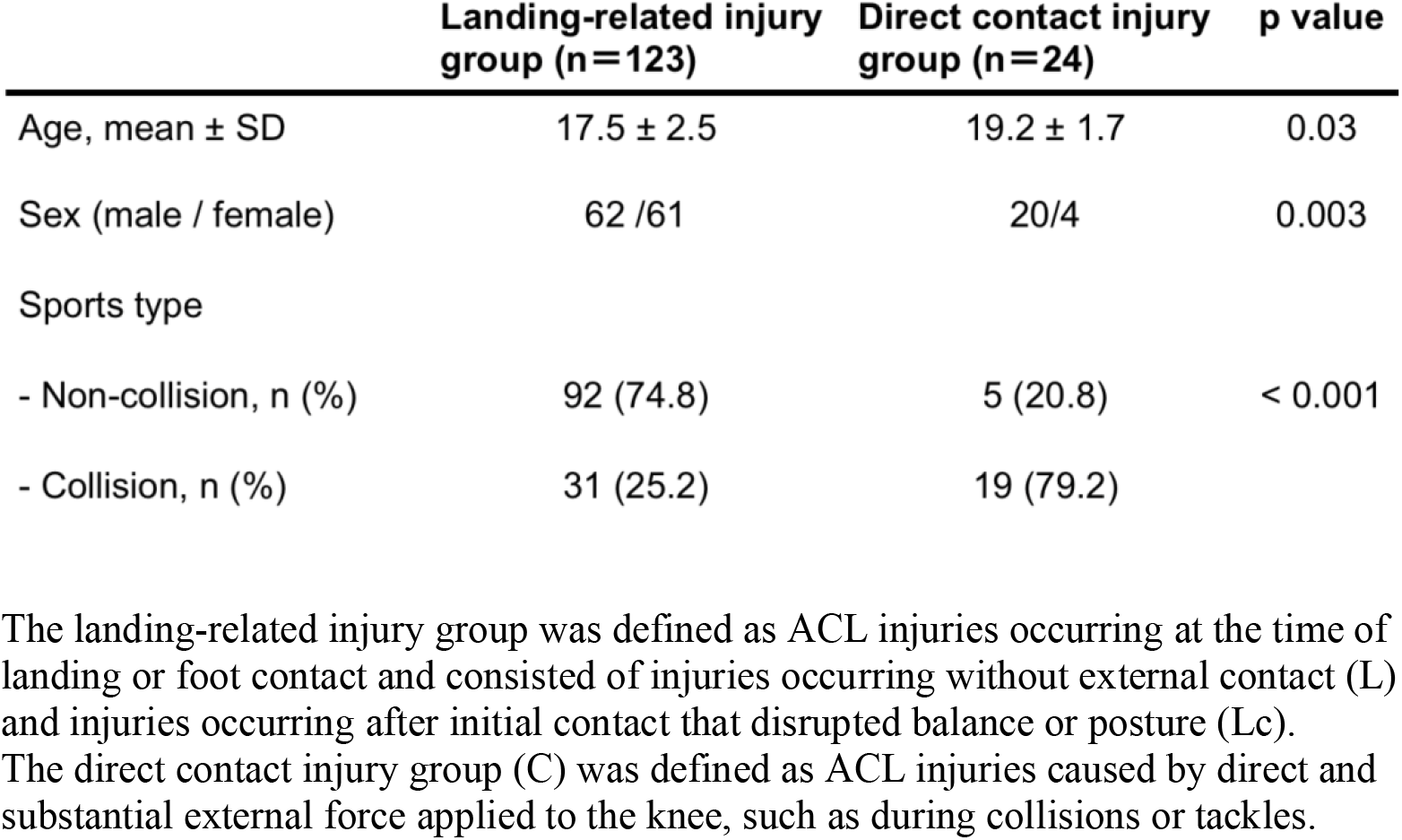
Comparison of demographic characteristics between landing-related injury group and direct contact injury group.

When injuries were analyzed according to sport contact characteristics, collision sports had a significantly higher proportion of direct contact injuries (Table 1).

### Multivariable Analysis

In the multivariable logistic regression analysis with landing-related injury (L + Lc) as the dependent variable, participation in noncollision sports was significantly associated with landing-related injury (odds ratio [OR], 9.80; 95% confidence interval [CI], 3.03– 31.5; p < 0.001).

Sex was not significantly associated with landing-related injury. Age was significantly associated with landing-related injury, with an OR of 0.71 per 1-year increase in age (Table 2).

**Table 2.** Multivariate logistic regression analysis for landing-related injury.

| **Variables** | **OR** | **95%CI** | **p value** |
| --- | --- | --- | --- |
| Age (per a year increase) | 0.71 | 0.52–0.97 | 0.034 |
| Female (vs male) | 1.17 | 0.29–4.68 | 0.83 |
| Non-collision sports (vs collision) | 9.80 | 3.03–31.5 | < 0.001 |

Landing-related injury (L + Lc) was used as the dependent variable. Independent variables included age, sex, and sport contact characteristics. OR, odds ratio; CI, confidence interval.

## Discussion

The most important finding of this study was that the majority of ACL injuries in athletes aged 22 years or younger occurred through landing-related injury mechanisms. Regardless of the presence or absence of external contact, landing-related injuries accounted for more than 80% of all cases, and non-collision sports were strongly associated with landing-related injury mechanisms. These findings suggest that the occurrence of ACL injury may be more closely related to insufficient control of landing and foot contact during sports movements than to the presence of direct external force applied to the knee itself.

Traditionally, ACL injuries have been broadly classified into contact and non-contact injuries. Non-contact injuries are defined as those occurring without direct external force to the knee, and intrinsic factors such as neuromuscular control deficits and abnormal alignment have been discussed as major contributors. ^1, 4, 8^ However, the results of the present study indicate that this dichotomous classification may not adequately reflect the actual injury mechanisms in young athletes. Landing-related injuries were observed not only in purely non-contact situations but also frequently during landing movements immediately following external contact. ^5–7^ The common biomechanical feature shared by these injury mechanisms was insufficient control of foot contact and landing during sports movements, regardless of whether contact was involved. ^5, 8^ These findings suggest that classifying ACL injuries solely based on the presence or absence of contact may fail to capture the mechanical processes leading to injury. A framework that re-evaluates ACL injury mechanisms from the perspective of landing and foot contact control may be more appropriate for understanding ACL injuries in young athletes. ^1, 4–7^

In this study, younger age was independently associated with landing-related injury. The decrease in the odds of landing-related injury with increasing age suggests that age-related differences in movement control may influence ACL injury mechanisms in young athletes. ^8–10^ Younger athletes may have immature neuromuscular control and a limited ability to absorb and regulate impact during landing. In addition, rapid changes in body size and muscle strength during growth may make it difficult to appropriately control foot placement and deceleration during sports movements. ^10^ As a result, landing-related injury mechanisms may occur even in situations without direct external force applied to the knee. ^4, 5, 8^ Importantly, these findings do not indicate that age itself is a direct cause of ACL injury. Rather, age may function as a surrogate marker reflecting developmental stage and neuromuscular control characteristics that influence the control of landing and foot contact. ^8–10^ Although female sex was more prevalent in the landing-related injury group in the unadjusted analysis, sex was not independently associated with landing-related injury after adjustment for age and sport contact characteristics. ^8, 9^

Collectively, these findings suggest that insufficient control of landing and foot contact represents a fundamental biomechanical process underlying ACL injury in young athletes, irrespective of whether external contact is present. ^1, 4–8^ Traditionally, ACL injury prevention strategies have often been organized around the presence or absence of collision or direct contact. However, the present findings indicate that a prevention framework centered primarily on landing and foot contact control may be more appropriate than one based solely on contact characteristics. ^1, 11^ Therefore, ACL injury prevention programs should emphasize the assessment and training of landing and foot contact control during sport-specific movements. Neuromuscular training targeting dynamic alignment, trunk stability, and deceleration control during landing may represent important interventions for young athletes, even in sports traditionally considered to involve minimal contact. ^8, 11, 12^

Overall, these findings indicate that ACL injury prevention should adopt a comprehensive approach across a wide range of sports, regardless of the degree of contact, with a primary focus on improving movement quality during landing and cutting maneuvers. ^1, 8, 11^

This study has several limitations. First, it was a retrospective, single-center study, which may limit the generalizability of the findings. Second, the classification of injury mechanisms was based on medical records and patient reports, and the possibility of recall bias and misclassification cannot be excluded. Third, detailed biomechanical data at the time of injury were not available, and causal relationships between landing control and ACL injury could not be directly examined. Nevertheless, the present findings demonstrate that the majority of ACL injuries in athletes aged 22 years or younger are associated with landing-related injury mechanisms, regardless of the presence or absence of contact. In addition, the association between younger age and landing-related injury suggests that developmental differences in movement control capacity may influence ACL injury mechanisms in young athletes. These findings support a comprehensive perspective that prioritizes the control of landing and foot contact during sports movements for the prevention of ACL injuries.

## Conclusion

In athletes aged 22 years or younger, the vast majority of ACL injuries occur through landing-related mechanisms, regardless of the presence or absence of external contact. These findings indicate that ACL injury mechanisms in young athletes are more closely associated with insufficient control of landing and foot contact than with direct external forces applied to the knee. A prevention framework centered on landing and foot contact control may therefore be more appropriate than the traditional contact versus noncontact classification when designing ACL injury prevention strategies for young athletes.

## Data Availability

The datasets generated and/or analyzed during the current study are not publicly available due to institutional privacy regulations but are available from the corresponding author on reasonable request.

## Acknowledgments

The authors thank the clinical staff of the sports injury clinic at Ashiya Central Hospital for their assistance with data collection.

## References

1. Griffin LY, Albohm MJ, Arendt EA, et al. Understanding and preventing noncontact anterior cruciate ligament injuries: a review of the Hunt Valley II meeting. Am J Sports Med. 2006;34(9):1512–1532.

2. Lohmander LS, Englund PM, Dahl LL, Roos EM. The long-term consequence of anterior cruciate ligament and meniscus injuries. Am J Sports Med. 2007;35(10):1756–1769.

3. von Porat A, Roos EM, Roos H. High prevalence of osteoarthritis 14 years after an anterior cruciate ligament tear in male soccer players. Acta Orthop Scand. 2004;75(1):75–82.

4. Boden BP, Dean GS, Feagin JA Jr, Garrett WE Jr. Mechanisms of anterior cruciate ligament injury. Orthopedics. 2000;23(6):573–578.

5. Krosshaug T, Nakamae A, Boden BP, et al. Mechanisms of anterior cruciate ligament injury in basketball: video analysis. Am J Sports Med. 2007;35(3):359– 367.

6. Waldén M, Krosshaug T, Bjørneboe J, Andersen TE, Faul O, Hägglund M. Three distinct mechanisms characterize anterior cruciate ligament injuries in professional football players. Br J Sports Med. 2015;49(22):1452–1457.

7. Olsen OE, Myklebust G, Engebretsen L, Bahr R. Injury mechanisms for anterior cruciate ligament injuries in team handball. Am J Sports Med. 2004;32(4):1002– 1012.

8. Hewett TE, Myer GD, Ford KR, et al. Biomechanical measures of neuromuscular control and valgus loading of the knee predict ACL injury risk. Am J Sports Med. 2005;33(4):492–501.

9. Myer GD, Ford KR, Khoury J, Hewett TE. Biomechanics laboratory-based prediction algorithm to identify female athletes at high risk for ACL injury. Am J Sports Med. 2010;38(10):2025–2033.

10. Quatman-Yates CC, Myer GD, Ford KR, Hewett TE. A longitudinal evaluation of maturational effects on lower extremity strength in female adolescent athletes. Pediatr Phys Ther. 2013;25(3):271–276.

11. Sugimoto D, Myer GD, Foss KD, Hewett TE. Specific exercise effects of preventive neuromuscular training intervention on anterior cruciate ligament injury risk reduction in young females. Am J Sports Med. 2015;43(11):2824– 2833.

12. Herman DC, Oñate JA, Weinhold PS, et al. The effects of feedback with and without strength training on lower extremity biomechanics. J Athl Train. 2009;44(2):123–130.

